# Bacterial quantification assay to detect and monitor *Pseudomonas aeruginosa* infection in patients with bronchiectasis

**DOI:** 10.1101/2025.10.03.25336435

**Authors:** Daniela Alferes de Lima Headley, Adam J Bell, Rebecca C Hull, Marie-Jeanne H C Kempen, Migle Young, Chandani Hennayake, Qingyou Du, Rachel Galloway, Eve McIntosh, Zsofia Eke, Hollian Richardson, Merete B Long, Sarah Williams-Macdonald, Hazel Buchanan, Edward Mountjoy, Mateja Sborchia, Pawlina Dand, Daniel L Halligan, James A Long, Maia Kavanagh Williamson, Georgina Vernon, Declan Fawcett-Gibson, Daniel De Vega, Oriol Sibila, Stefano Aliberti, Charles S Haworth, Rachel Dakin, Rebecca Holmes, Carolyn Clarke, James D Chalmers

## Abstract

**Background:** *Pseudomonas aeruginosa* (PA) is the most commonly detected pathogen in bronchiectasis. The clinical standard of care for pathogen detection is culture, which has low sensitivity. Early detection and improved monitoring could be used to enhance eradication and long-term suppressive treatments.

**Methods:** A novel bacterial quantification assay (BQA) targeting ribosomal RNA for the detection of viable PA was developed. The BQA sensitivity and specificity in patient samples was assessed utilising culture (n=57); as well as BioFire pneumonia panel (n=111) and 16S rRNA gene sequencing (n=62). The potential clinical impact of the BQA in clinical settings was explored using two international patient cohorts.

**Results:** The BQA detected PA in 100% of samples where PA was identified by other methods. The BQA quantification was positively correlated with quantitative culture (r=0.42; p<0.001) and BioFire (r=0.68; p<0.001). The BQA identified PA in an additional 23-44% of cases. To evaluate if this increased detection had clinical implications, we tested sputum samples considered to be PA negative by culture in patients who subsequently tested positive for PA. The BQA detected PA in 8/20 patient samples 8-484 days prior to primary detection by culture. In patients considered to have successful PA eradication by culture, the BQA detected PA in 13 samples from 27 patients who went on to relapse.

**Conclusions:** The BQA is a highly sensitive detection and quantification method for PA. The BQA demonstrated improved detection and treatment monitoring compared to culture, identifying patients who went on to either isolate a first PA or relapse.

## Introduction

Bronchiectasis is a chronic lung disease characterised by irreversible enlargement of the bronchi, inflammation, infection and airway dysfunction (1–3). Patient symptoms include sputum production, fatigue, chronic cough and recurrent respiratory infections (4, 5). Bronchiectasis was initially described as a rare disease, however recent decades have seen an increase in diagnosis worldwide, with reported rates between 53-1,249 per 100,000 individuals dependent on geographical location (6, 7).

A challenging aspect of disease management is recurrent and chronic respiratory infections. The most common pathogens detected in patient samples are *Pseudomonas aeruginosa*, *Haemophilus influenzae, Enterobacteriaceae, Staphylococcus aureus, Streptococcus pneumoniae* and *Moraxella catarrhalis* (4). The prevalence of each pathogen is dependent on geographical location, with *P. aeruginosa* being the most common overall (4). *P. aeruginosa* is of particular clinical importance as it is associated with increased exacerbation frequency, risk of hospitalisation and mortality (8, 9).

Clinical care for patients with bronchiectasis is guided exclusively by traditional bacterial culture methods (10, 11). Culture-based methods, however, have low sensitivity and may not detect organisms present at low bacterial loads or otherwise viable but non-culturable (VBNC) organisms present in patient samples (10–12). VBNC organisms lose the ability to grow on routine media but remain metabolically and transcriptionally active allowing their detection through non-culture based methods including reverse transcriptase quantitative PCR (RT-qPCR) (12). Recently, molecular based assays have been developed to rapidly identify pathogens. Molecular detection may increase the sensitivity for bacterial detection from samples such as sputum. In a recent study in patients with bronchiectasis a multiplex platform called the BioFire Film Array Pneumonia Plus Panel increased the detection of bacterial pathogens at exacerbation in 120 patients with bronchiectasis from 68% detected by culture to 86% detected using PCR. This assay utilises a nested multiplex polymerase chain reaction (PCR) approach targeting nucleic acids (DNA and RNA) to detect 9 viruses and 18 bacteria and 7 antimicrobial genes (13).

European Respiratory Society guidelines recommend early eradication of *P. aeruginosa* with antibiotic therapy, emphasizing the importance of detecting this bacteria early and accurately (14). Reported success of eradication therapy in bronchiectasis is around 40%. This is lower than is reported in cystic fibrosis (60-90%), which may be attributed to later diagnosis and less frequent monitoring for new infections. *P. aeruginosa* detection changes clinical practice, with different antibiotic management at exacerbation, and a different algorithm for treatment involving inhaled antibiotics in patients with frequent exacerbations (15, 16). Consequently, a test that could identify *P. aeruginosa* (in sputum) earlier in the disease course, and with higher sensitivity than culture (the current clinical standard of care), would be of high clinical value.

The BioFire Film Array Pneumonia Plus Panel is designed for DNA and RNA detection and may not be suitable for use in treatment monitoring or in patients receiving antibiotics, because it may not distinguish between viable and non-viable organisms. (17–19). An attractive target that has shown correlation with viability is ribosomal RNA (rRNA) (20, 21). There is precedent for the use of rRNA for the detection and quantification of viable bacteria such as for the accurate detection and quantification *Mycobacterium tuberculosis* in sputum. (22, 23). This assay has shown great promise for monitoring *M. tuberculosis* infections, with comparable performance to current gold standards for diagnosis and greater accuracy than culture for monitoring treatment response (22).

Here we describe the clinical utility of a newly developed bacterial quantification assay (BQA) targeting rRNA for the detection of viable *P. aeruginosa* from clinical samples in bronchiectasis patients.

## Methods

### Patient Cohorts

Patient cohorts were used to address three clinical questions: Firstly, how detection of *P. aeruginosa* using the BQA compared to culture, and DNA based molecular based pathogen detection. Secondly, based on higher sensitivity of molecular testing compared to culture, whether *P. aeruginosa* infection could be identified earlier in patients with bronchiectasis prior to their first positive culture. Finally, to evaluate whether the BQA could be used to monitor response to treatment we used samples from patients enrolled in a randomized controlled trial of an inhaled antibiotic treatment to determine if the BQA could identify clearance of *P. aeruginosa* (referred to as eradication) and detect early relapse.

To answer these questions sputum samples were obtained from two bronchiectasis patient cohorts. For both cohorts, sputum was obtained from participants and stored at –80°C prior to processing.

The EMBARC-BRIDGE study is a multicentre prospective observational study embedded within the European bronchiectasis registry (4, 24, 25). Patients enrolled provided sputum samples at a baseline visit and used for molecular microbiology. Where sufficient sample was available, sputum from the same clinical visit was sent for routine clinical microbiology to compare culture results with the results of the molecular assays, and 16S rRNA gene sequencing was performed (as described below). The BRIDGE study is registered under ClinicalTrials.gov ID: NCT03791086.

The ORBIT 3 and 4 trials were double blind randomized placebo-controlled trials of inhaled liposomal ciprofloxacin versus placebo in patients with chronic *P. aeruginosa* infection (26). Sputum samples at monthly study visits were sent to the central lab for determination of bacterial load and detection of *P.* aeruginosa by quantitative culture. Samples were stored at –80°C and then transferred to the EMBARC bioresource for further analysis at the end of the trial. Specific samples were selected for analysis of treatment response by identifying patients who had achieved “eradication”: defined as 3 or more negative sputum samples following treatment, and no subsequent positive sputum samples for *P. aeruginosa* by culture. “Relapse” was defined as a minimum of two consecutive negative sputum samples followed by a positive sample by culture. The ORBIT clinical trials were registered under ClinicalTrials.gov IDs: NCT01515007and NCT02104245.

### RNA extraction

Sputum RNA was extracted using the ZymoBIOMICS RNA Miniprep kit (Zymo Research Cat. No. R2001) according to manufacturers’ instructions with small modifications. Full details are described in the supplement methods.

### Dual nucleic acid extraction

Sputum RNA and DNA were extracted simultaneously using the AllPrep DNA/RNA Mini kit (Qiagen Cat. No. 80204) according to manufacturers’ instructions with small modifications. Full details are described in the supplement methods.

### RT-qPCR

The clinical utility of the BQA was evaluated in collaboration between LifeArc and the University of Dundee.

One-step RT-qPCR was performed to quantify *P. aeruginosa* rRNA from patient sputum samples. The general reaction mix per reaction used 1x master mix, 1x reverse transcriptase enzyme, optimised concentration of primers and dual labelled probe (IDT) with water making up the total reaction volume to 8µl. The dual labelled probe was tagged with a 5’ 6-FAM reporter dye and a 3’ non-fluorescent quencher. A DNA contamination control mastermix (RT negative) was also prepared, to ensure rRNA quantification was reliable. The RT negative reaction mix was identical to the general mix except for absence of reverse transcriptase enzyme.

RT-qPCR was performed using a QuantStudio 7 Flex Real-Time PCR machine (Applied BioSystems). 2µl of a one in ten dilution from each sample was used per reaction. Samples, positive control and water controls were tested in triplicates. Samples with a quantification of ≥4.57 copies/reaction were considered positive for *P. aeruginosa.* Cut-off rationale explained in supplement methods.

Standard curves were performed as required, with 1 in 10 serial dilutions prepared from a known concentration positive control to obtain copies per reaction for each sample.

### Multiplex PCR (BioFire)

Respiratory pathogens from sputum samples were identified by a nested multiplex PCR, BioFire Film Array Pneumonia Plus Panel with the FilmArray 2.0 multiplex PCR system (Biomerieux) as previously described (27).

### 16S rRNA gene sequencing (16S)

Sputum DNA was extracted using the DNeasy PowerSoil Pro Kit (Cat. No. 47014). DNA was sequenced by synthetic full-length 16S rRNA gene LoopSeq (28). Reads quality was checked using FASTQC and MultiQC in R 4.1.2 (2022-12-30). ASVs were generated through DADA2 pipeline with taxonomy assignment to species level utilising Silva v.138.1 database (29). Taxa identified were reported as a positive result if present with more than ten reads or with >0.1% relative abundance in a sample. Sequencing data have been submitted to European Nucleotide Archive (ENA) (Project accession number PRJEB88963).

### Statistical analysis

Sensitivity and specificity were calculated as previously reported (30). 95% confidence intervals (CI) were calculated utilising the percentile method using bootstrapping with 500 iterations. Correlation analyses were performed using either Spearman’s rank or Pearson’s correlation coefficient as appropriate.

## Results

### Analytical performance of the BQA

The analytical performance of the *P. aeruginosa* specific rRNA targeting RT-qPCR assay was confirmed (Supplement methods). The BQA has a wide linear interval of 42 – 4.95×10^7^ copies/reaction, with low limit of quantification (LOQ) and limit of detection (LOD) of ≥688 and 40 copies/reaction, respectively. The assay was highly reproducible, with covariance of <10% for intra-assay precision and repeatability between assay days (Supplement methods and Table S1).

### Comparison of the BQA with molecular methods and culture

To demonstrate the suitability of the BQA to identify *P. aeruginosa* in patient sputum, 20 samples from the ORBIT clinical trials with culture-confirmed *P. aeruginosa* infection (bacterial load range of 1.6-9 log CFU/g), were tested. 100% concordance was found with the culture positive samples and BQA results (20/20 were positive by BQA).

The performance of the BQA was compared with BioFire, culture and 16S rRNA gene sequencing. 111 samples obtained from 98 bronchiectasis patients as part of the EMBARC BRIDGE study were used in this investigation. 54 (55.1% of patients were male), with a mean age of 64.8 years (standard deviation 14.1). The mean BMI was 25.9 (SD 5.5) and FEV1% predicted was 75.6% (SD 26.3). 41 (41.8%) patients had at least 2 exacerbations in the previous year. 38 (38.8%) patients had idiopathic bronchiectasis, 15 (15.3%) had post-infective bronchiectasis, 12 (12.2%) COPD, 9 (9.2%) primary ciliary dyskinesia, 7 (7.1%) asthma and 5 (5.1%) NTM. Other causes of bronchiectasis accounted for <4 cases. All 111 samples were processed for both BioFire and the BQA; of these 57 had previous culture results and 62 had sequencing data available.

Compared to BioFire, the BQA demonstrated a sensitivity of 100% (24/24; 95% CI: 100 – 100%) and a specificity of 74.7% (65/87; 95% CI: 64.4 – 83.4%), indicating that the BQA identified *P. aeruginosa* in 22 BioFire negative samples. Compared to culture, the BQA demonstrated a sensitivity of 100% (5/5; 95% CI: 100 – 100) and a specificity of 63.5% (33/52; 95% CI: 50.4 – 77.6), indicating that the BQA identified *P. aeruginosa* in 23 culture negative samples. Compared to 16S rRNA gene sequencing, the BQA demonstrated a sensitivity of 100% (14/14; 95% CI: 100 – 100) and a specificity of 81.2% (39/48; 95% CI: 69.3 – 92.1), indicating that the BQA identified *P. aeruginosa* in 9 sequencing negative samples (Figure 1).

**Figure 1.**
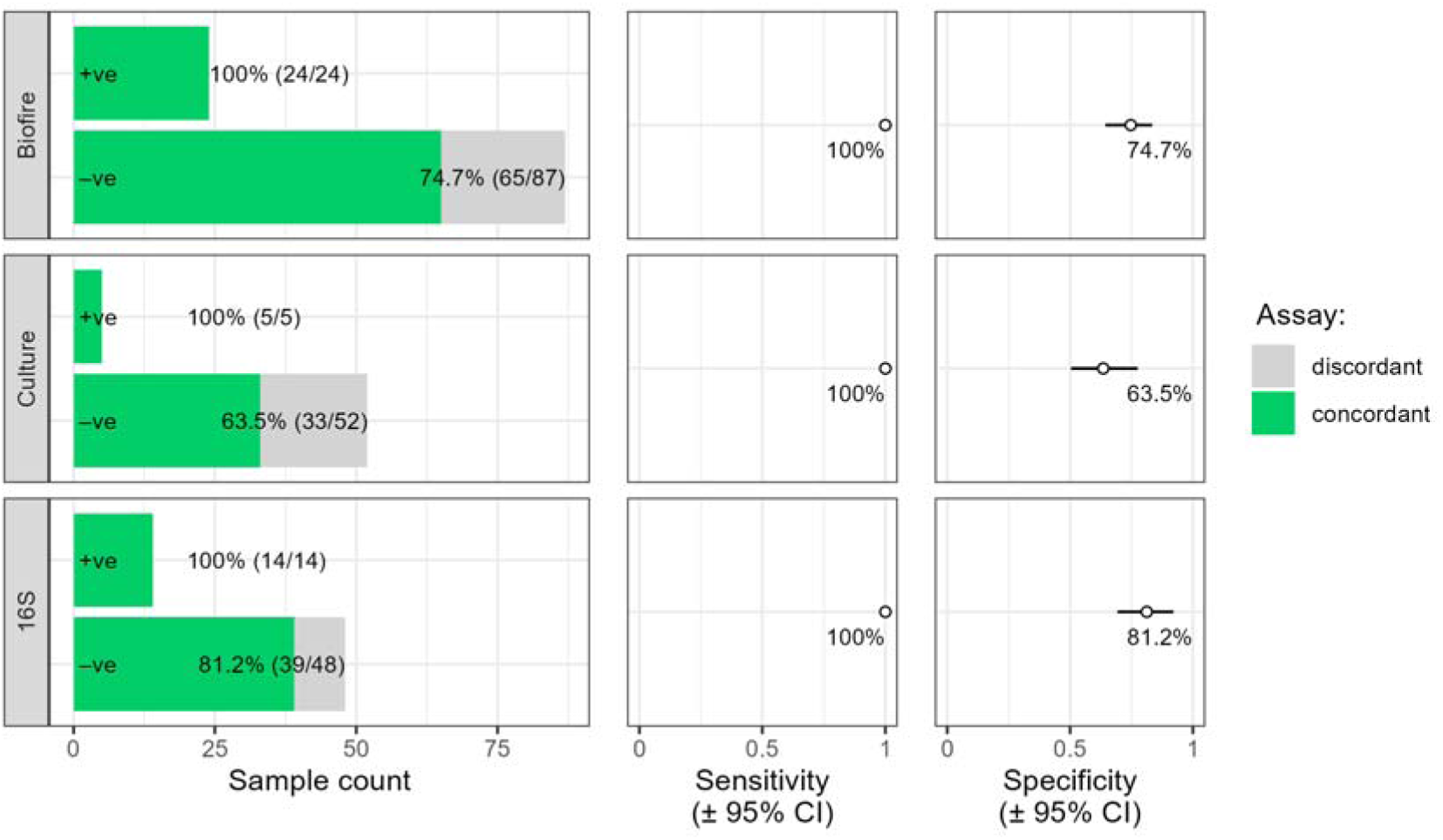
Sensitivity and specificity of the bacterial quantification assay when compared to the BioFire Pneumonia Plus Panel, Culture and 16S rRNA gene sequencing status.

Neither culture nor DNA targeting methods can be considered a true gold standard, due to low sensitivity and inability to distinguish between viable and non-viable organisms respectively; therefore, it is difficult to establish whether the BQA is identifying clinically important *P. aeruginosa* which is missed by the other methods, or whether BQA identifies false positives.

### The BQA quantification correlates with BioFire semi-quantification and quantitative culture

BioFire provides a semi-quantifiable result, to investigate the relationship with the BQA quantification we performed a correlation analysis (Figure 2). A strong positive correlation between BioFire and BQA quantification was observed (Spearman’s ρ = 0.68, 95% CI: 0.57 – 0.77, p < 0.001).

**Figure 2.**
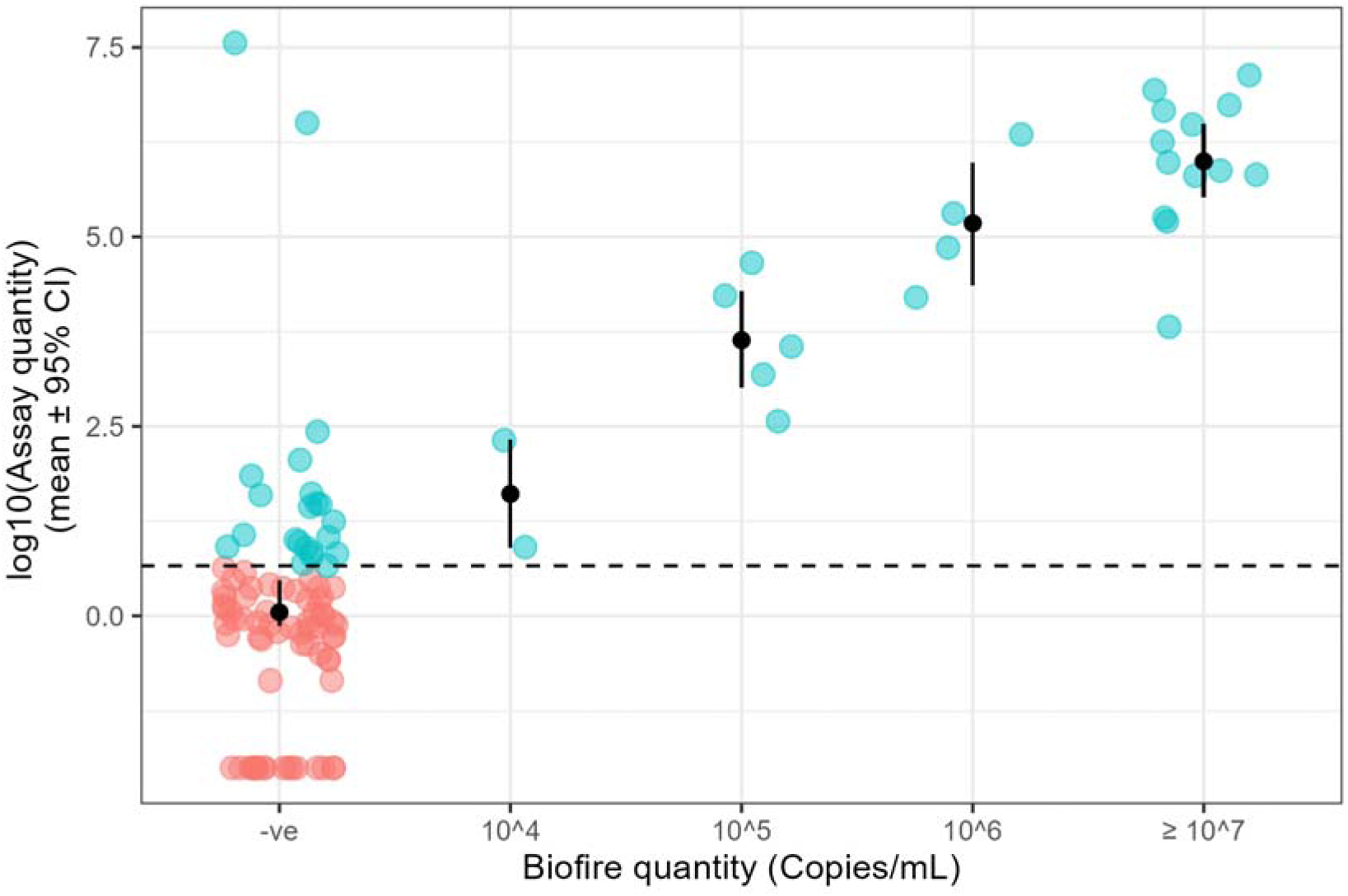
Comparison of BioFire and the bacterial quantification assay quantitative outputs (n = 111). Each point represents a single sputum sample, and compares the results for both the bacterial quantification assay and BioFire. Hashed line represents the bacterial quantification assay cut off (4.57 copies/reaction), with red points representi negative BQA samples and blue points represent positive BQA samples.

To determine the relationship of the BQA quantification with quantitative culture (CFU/g), 113 baseline samples from the ORBIT trials (*P. aeruginosa* culture positive) were used and a correlation analysis performed (Figure 3). A moderate positive correlation was observed between quantitative culture and BQA quantification (Pearson’s r = 0.42, CI: 0.25 – 0.56, p < 0.001).

**Figure 3.**
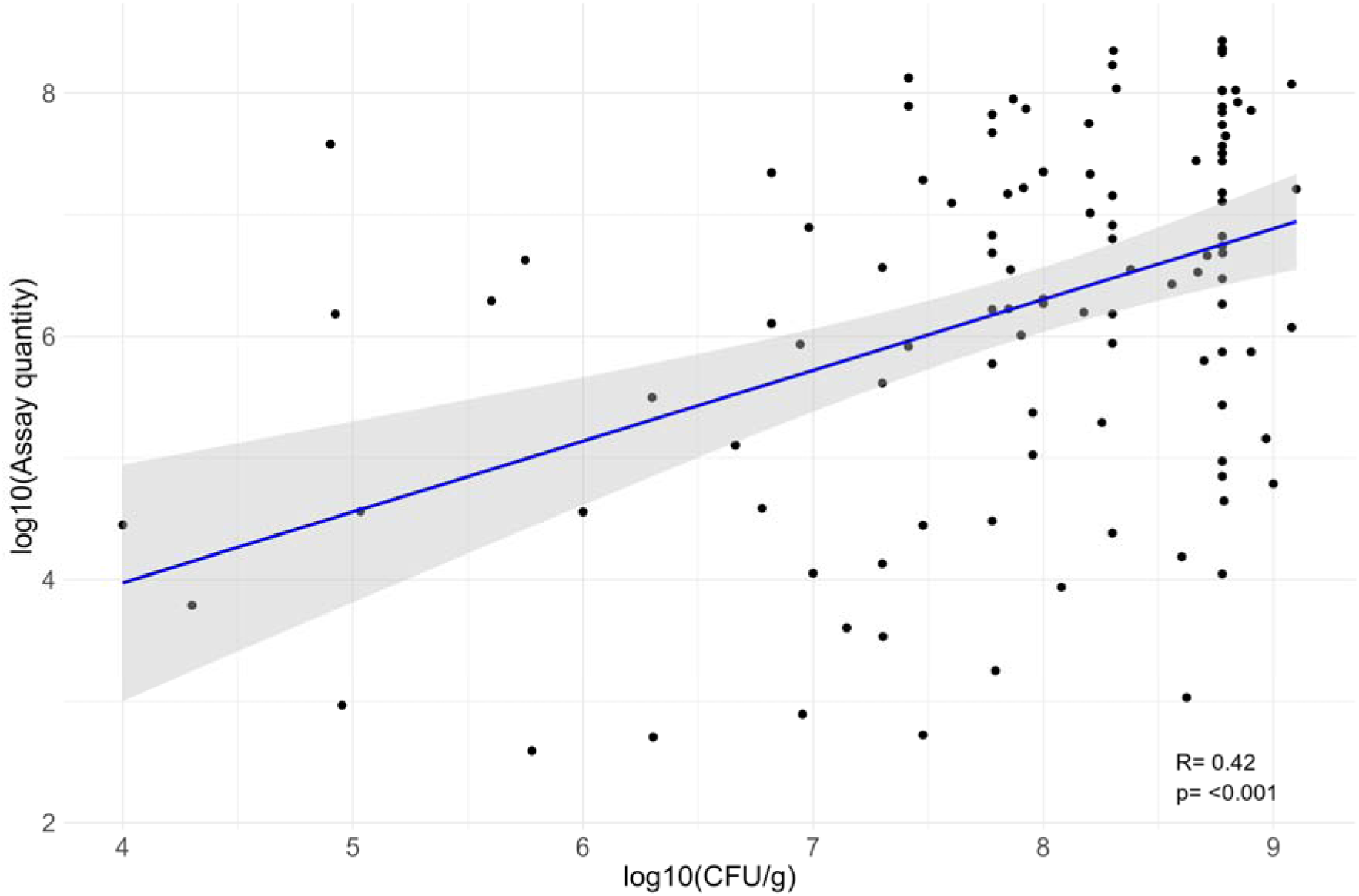
Comparison of quantitative culture (CFU/g) and the bacterial quantification assay quantitative outputs (n = 113). Each point represents a single sputum sample, and the results for the BQA and quantitative culture. Blue line represents regression line, with 95% confidence interval shown in grey.

### The BQA detects *P. aeruginosa* prior to culture positivity

A benefit of molecular based methods for pathogen detection is their higher sensitivity and their ability to detect organisms that are VBNC. As such using a molecular based method might allow earlier detection of pathogens than culture.

15 patients were identified who had a first or new isolation of *P. aeruginosa* during the BRIDGE study. Sputum samples which were negative for *P. aeruginosa* and obtained 8 to 484 days prior to primary *P. aeruginosa* detection by culture were then studied to identify if the BQA could identify *P. aeruginosa* earlier than culture.

8/20 (40.0%; exact 95% CI: 19.1 – 63.9%) samples from 7/15 patients were positive by BQA for *P. aeruginosa* rRNA (6.75 x 10^2^ – 7.28 x 10^6^ copies/reaction) (Figure 4). The mean days prior to culture detection were 138 (range of 41-298). Full analysis details can be found in Supplement Table S2. This provides preliminary evidence of the potential for the BQA to detect *P. aeruginosa* at an earlier timepoint than culture-based methods in some patients.

**Figure 4.**
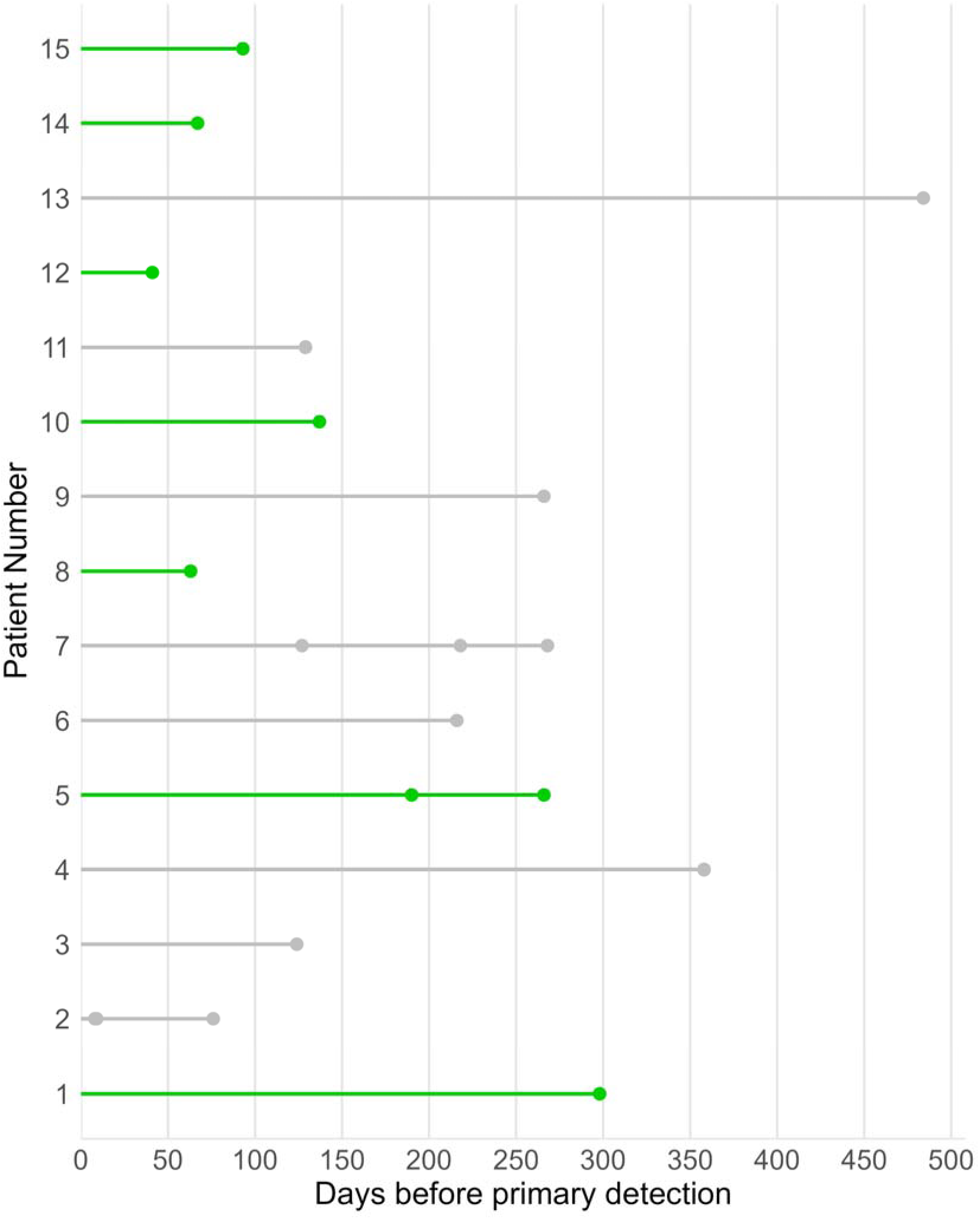
P. aeruginosa bacterial quantification assay status of each sample (per patient) and the number of days before primary *P. aeruginosa* detection by culture. Green indicates samples positive by the bacterial quantification assay, and grey negative.

### The BQA shows potential as a treatment monitoring tool

To explore the potential of the BQA for infection treatment monitoring, 36 patients were selected from the ORBIT clinical trials based on *P. aeruginosa* infection status. Infection status was designated as eradication (n=10) or relapse (n=27, one patient had two relapses). Eradication was defined by 3 consecutive negative sputum cultures, and relapse, by a *P. aeruginosa* positive sample following at least 2 consecutive negative samples. The BQA quantification was compared to quantitative culture (CFU/g) for concordance.

For the relapse cohort two culture negative samples (first available culture negative and final available culture negative prior to relapse) and the positive sample indicating relapse were selected (where available). 13/20 (65.0%; exact 95% CI: 40.8 – 84.6%) ‘first culture negative’ samples were positive by BQA (Figure 5). The trend continued with the ‘final available culture negative’ samples with 14/23 (60.9%; exact 95% CI: 38.5 – 80.3%) positive (Figure 5). Lastly, 14/15(93.3%; exact 95% CI: 68.1 – 99.8%) of relapse samples were positive, with the one negative sample having a 1.6 log CFU/g by culture (Figure 5).

**Figure 5.**
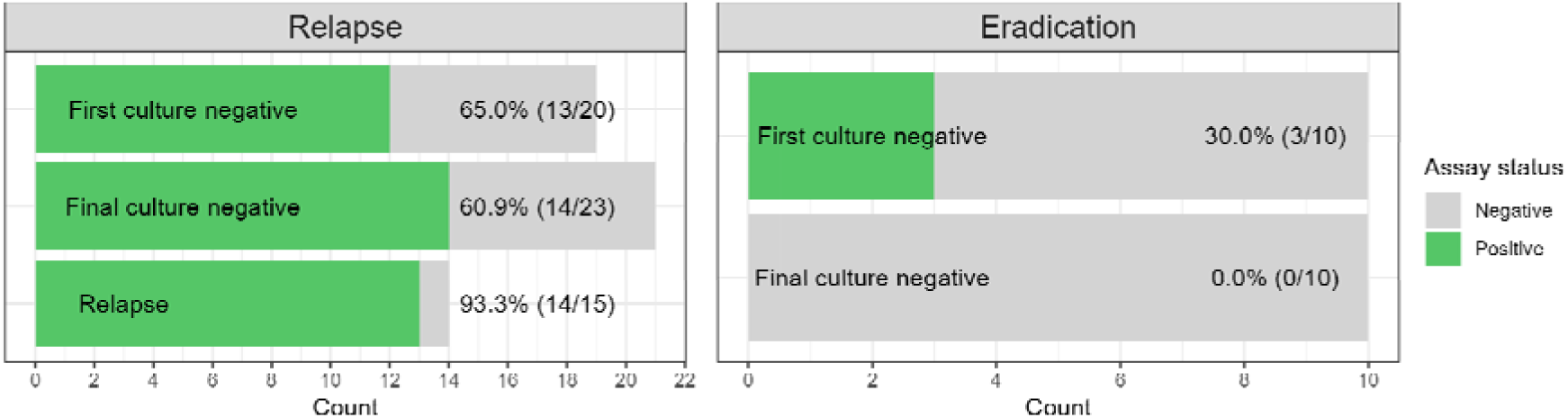
P. aeruginosa bacterial quantification assay status for samples from patients who went onto a P. aeruginosa relapse or eradication with relation to their culture status. In red are bacterial quantification assay negative results and in green bacterial quantification assay positive results.

**Figure 6.**
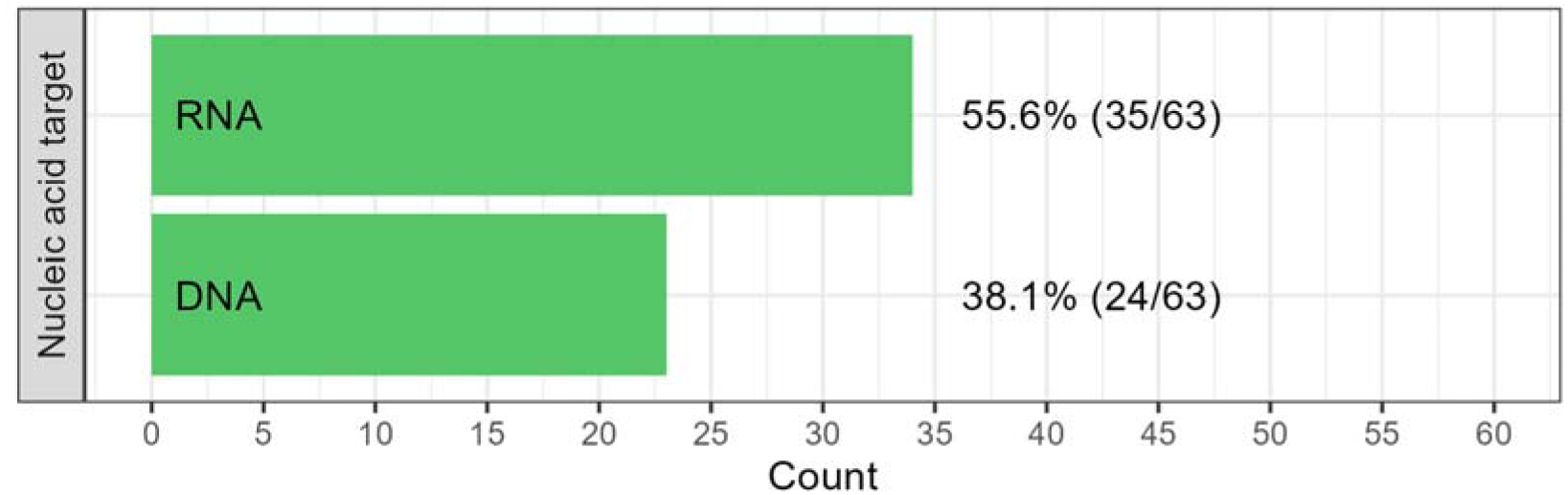
P. aeruginosa bacterial quantification assay positive status count using rRNA and DNA as assay targets for samples from patients who were P. aeruginosa culture negative, but went onto have P. aeruginosa detected via culture at a later date.

For the eradication cohort two culture negative samples (first available culture negative and further culture negative prior to end of clinical trial) were selected. 3/10 (30%; exact 95% CI: 6.7–65.2%) ‘first culture negative’ samples were positive, with no positive samples at a later timepoint (Figure 5). Full analysis details can be found in Supplement Table S3.

### rRNA is more sensitive than DNA at detecting *P. aeruginosa* from patient samples

As most current molecular methods used for diagnostics target DNA, we compared the sensitivity of rRNA and DNA as targets for the BQA. Dual nucleic acid extractions were performed on samples with *P. aeruginosa* culture negative status from patients who went onto receive a positive culture status at a later date. Both DNA and rRNA were used as targets for the BQA and their *P. aeruginosa* detection percentage was compared. Detection of *P. aeruginosa* was significantly higher when targeting rRNA compared to DNA (55.6% vs 38.1%; difference = 17.5%, 95% CI: 0.3 – 34.6%; p = 0.0495). rRNA quantification had a median of 1.04×10^3^ copies/reaction (range of 5.67 – 4.73×10^6^) and DNA quantification had a median of 3.73×10^2^ copies/reaction (range of 9.98 – 4.10×10^4^). Full analysis details can be found in Supplement Tables S2 and S3.

## Discussion

We have described the clinical utility of a newly developed BQA for the detection of viable *P. aeruginosa* from bronchiectasis patient samples. The main goal was to determine the suitability of the BQA in a clinical setting. To do so we compared the assay performance with the current clinical standard of care (culture), and two additional methods (16S rRNA gene sequencing and the molecular based assay BioFire Film Array Pneumonia Plus Panel). The BQA showed high sensitivity when compared to all three methods (100%), and also identified a high proportion of additional positive samples which were not identified by these other methods. This could indicate either superiority of the BQA, or a lack of specificity (“false positives”). A major challenge in the development of a novel assay like this is that there is no true gold standard, since culture is known to have poor sensitivity. As such, it is not certain that all of the additional positive *P.* aeruginosa detections with the BQA are clinically significant. We therefore took advantage of the availability of unique well characterised patient cohorts to explore scenarios where the BQA could be used clinically. Several findings from testing these samples indicate that positive detection of *P. aeruginosa* using BQA when other assays tested negative is due to increased sensitivity of this assay and not decreased specificity. First, other identification that there were no “false positives” using the BQA at late timepoints after successful eradication makes it less likely that the BQA has lower specificity than the other assays. Second, the superior sensitivity compared to the other assays in patients who subsequently relapsed (tested positive by culture) supports the conclusion that the BQA is more sensitive but remains highly specific. An additional benefit of the BQA is quantification, which can be useful for assessing clinical significance and monitoring treatment response. The BQA quantification showed a strong positive relationship with BioFire quantification.

Alongside higher sensitivity compared to culture; it has been shown that molecular methods can detect VBNC. It is believed that initial infection with *P. aeruginosa* is likely to be preceded by either prior infections with *P. aeruginosa* that are cleared, or low-level *P. aeruginosa* infection that may evade detection until it comes to dominate the microbiome (31). Therefore, there is theoretically an opportunity for molecular methods to identify *P. aeruginosa* infection prior to the first culture positive. Here we showed that the BQA detected *P. aeruginosa* up to 298 days prior to primary detection by culture. The data from the EMBARC cohort are limited by the variable timepoints at which the prior sputum samples were obtained as part of routine clinical practice. Therefore, the possibility of a new infection developing between BQA positive and culture positive timepoints cannot be fully excluded. Nevertheless, the ability to detect *P. aeruginosa* infection earlier was confirmed by the “relapse” analysis in the ORBIT trials where monthly samples prior to a positive sample showed that culture negative samples were positive earlier with the BQA. It is not possible to definitively ascertain whether the BQA positivity is due to higher sensitivity compared to culture or due to detection of VBNC however as the range of copies/reaction was 5.67 – 4.73×10^6^, it would suggest that positive results may be due a combination of these. In a clinical setting earlier detection may have allowed treatment to begin sooner. It is believed that eradication treatment, as recommended by ERS guidelines, is likely to be more effective if started closer to the date of first infection (32). Although both nucleic acids were positive during culture negative states using the BQA, rRNA was shown to be more sensitive than DNA detecting a higher number of positive samples, highlighting a potential benefit of using rRNA as the target for this BQA assay.

The BQA detected *P. aeruginosa* in culture negative samples prior to infection relapse. This is unlikely to be due to rRNA survival following cell death as for 3 of the patients the second culture negative sample had a higher copy number than the first with a time gap of 4 to 24 weeks between samples. Furthermore, no further culture negative samples from patients who went onto *P. aeruginosa* eradication were BQA positive, with a time gap of 4 to 40 weeks between samples. This is an initial indication of the BQA’s ability as a treatment monitoring tool, but further validation is required due to small patient numbers. Patients with three consecutive negative samples were chosen as the most likely to be genuine eradications. The lack of positive results in these samples supports that genuine false positive results with the BQA are unlikely.

Apart from utility in a clinical setting, another potential BQA application is clinical trials where monitoring the efficacy of treatments is crucial. Current methods to monitor bacterial infection during clinical trials is culture based, this is labour intensive and requires processing of samples in a timely manner which either requires the rapid shipment of samples to a central facility which is costly or processing at different centres which may lead to inconsistencies (26). In comparison, if the BQA is shown as a suitable alternative, samples could be appropriately stored and processed at the end of trial with a substantial cost saving. This should be evaluated in future studies.

We conducted initial investigation and assessment of the assay in small subsets of patients with specific characteristics, but larger scale clinical testing of the assay is now required in unselected patient groups.

In conclusion, we have developed and evaluated a novel BQA for the detection of viable *P. aeruginosa* from bronchiectasis patient samples. The BQA has shown comparability with current methods used in a clinical setting, with the potential to detect *P. aeruginosa* prior to primary culture identification and to detect *P. aeruginosa* in culture negative samples prior to an infection relapse. The BQA results demonstrate potential for clinical application in early diagnosis and treatment monitoring.

## Data Availability

All data produced in the present work are contained in the manuscript

## Acknowledgements

We acknowledge all of the members of the EMBARC Clinical Research Collaboration and the European Lung Foundation/EMBARC patient advisory group.

## Ethics statement

The study was approved by the research ethics committee in each participating country (18/LO/1935) and all patients gave written informed consent to participate.

## Support statement

Development and validation of the BQA was funded by LifeArc. This study is funded by the European Respiratory Society through the EMBARC3 Clinical Research Collaboration. EMBARC3 is supported by project partners Armata, AstraZeneca, Boehringer Ingelheim, Chiesi, CSL Behring, Glaxosmithkline, Grifols, Insmed, LifeArc, Roche, Verona and Zambon. JDC is supported by the GSK/Asthma and Lung UK Chair of Respiratory Research.

## Conflict of interest

JDC reports grants awarded from AstraZeneca, Boehringer Ingelheim, Chiesi, Genentech, Gilead, Grifols, Insmed and Trudell; consulting fees received from Antabio, AstraZeneca, Boehringer Ingelheim, Chiesi, Glaxosmithkline, Grifols, Insmed, Janssen, Novartis, Pfizer and Zambon. CSH has received research funding from AstraZeneca and fees for consultancy work and / or educational talks from 30 Technology, AstraZeneca, BiomX, Boehringer-Ingelheim, Chiesi, Clarametyx, Infex, Insmed, LifeArc, Pneumagen, Sanofi, Vertex and Zambon. OS has received payment or honoraria for lectures, presentations, speakers bureaus, manuscript writing or educational events from Insmed and B-I. SA reports grants or contracts from Glaxosmithkline, and consulting fees from Insmed Ireland Limited, Fondazione Internazionale Menarini, Insmed Italy S.R.L., Brahms Gmbh, Modernatx, Inc, Moderna Italy Srl, An2 Therapeutics, Inc, Physioassist Sas, Vertex Pharmaceuticals (Europe) Limited, Zambon Italia S.R.L., Zambon S.P.A., Chiesi Farmaceutici Spa, Insmed Germany Gmbh, Insmed Netherlands B.V, Boehringer Ingelheim Italia Spa, Insmed Incorporated, Boehringer Ingelheim International Gmbh, Sanofi S.R.L., Glaxosmithkline Spa. All other authors reported no conflicts of interest.

## Supplementary methods

### RNA extraction from sputum

In brief, 0.1g of sputum was homogenised in ZR bashing bead lysis tubes with DNA/RNA shield (1000µl of 1x or 650 µl of 2x concentration) for 40 seconds with a 15 second pause at 6200rpm using a Precellys instrument. 2 volumes of RNA lysis buffer were added to the supernatant, the full sample was then processed according to manufacturers’ instructions with DNAse I treatment.

For RNA extractions where *Staphylococcus aureus* was suspected, a lysostaphin step was performed prior to mechanical lysis. The addition of lysostaphin was shown to not impact *P. aeruginosa* yield (unpublished data).

### Dual nucleic acid extraction

In brief, 0.1g of sputum was homogenised in ZR bashing bead lysis tubes with 600µl of Buffer RLT plus with β-Mercaptoethanol for 40 seconds with a 15 second pause at 6200rpm using a Precellys instrument. The full sample was then processed according to manufacturers’ instructions. The RNA sample was DNAse I treated with RNA cleanup using the RNeasy Mini kit following manufacturers’ instructions (Qiagen Cat. No. 74104).

### RT-qPCR background amplification cut-off

To determine *P. aeruginosa* positive cut-off, the average quantification for Cq 35 of 14 plates was used. This provided us with 4.57 copies/reaction as our cut-off. Cq 35 cut-off reliably differentiated between true and background amplification during assay development activities. Cq 35 had been determined as a cut-off for background amplification during assay development (data not shown).

### Bacterial culture

*P. aeruginosa* (NCTC 12903) cultures were grown at 37°C in LB Miller broth, with an initial overnight culture followed by a subculture in fresh LB Miller broth for 4 hours.

### RNA extraction from bacterial culture

RNA was extracted as described above with small modifications. 1.5ml of pelleted *P. aeruginosa* culture resuspended in 350 µl residual media/PBS was homogenised with 700 µl of DNA/RNA shield in ZR bashing bead lysis tubes for 40 seconds at 6000rpm using a Precellys instrument. The remainder of the extraction was performed as previously described.

### Prototype assay analytical performance

The prototype assay analytical performance was evaluated. Template consisted of *P. aeruginosa* total RNA extracted from culture. Quantities for RNA were determined using standard curves generated from a 10-fold dilution series of a positive control at known concentrations (100 – 1×10^7^ copies/reaction). Key performance characteristics of intra-assay precision, repeatability, limit of detection (LOD), limit of quantification (LOQ), and linearity were assessed. Where available methods were adapted from relevant Clinical Laboratory Standards Institute (CLSI) standards.

Intra-assay precision was assessed in single RT-qPCR runs performed by operators paired with either a QuantStudio 6 Real-Time PCR System (QS6) or a Quant Studio 7 Flex Real-Time PCR System (QS7). Total RNA with rRNA concentrations of approximately 1×10^6^ copies/reaction (High), 1×10^4^copies/reaction (Medium), and 1×10^3^ copies/reaction (Low) were tested up to 42 times each. The percentage coefficient of variation (%CV) in reported quantities was used as the measure of intra-assay precision.

Methods of assessing repeatability were adapted from CLSI EP05A-A3 (1). The assessment followed a 20 x 4 x 1 manner, with testing across 20 days, 1 run per day, with 4 replicates per run of total RNA at High, Medium, and Low rRNA concentrations.

The LOD and LOQ were assessed across a 2-fold dilution series of total RNA with rRNA concentrations of 21-2.2×10^4^ copies/reaction. Data was collected from 90 replicates per concentration. Data analysis was performed using the method reported previously (2), which are similar to the probit approach outlined in CLIS EP17-A2 (3). The LOD was defined as the concentration predicted to yield 95% positive reactions. The LOQ was defined as the lowest assessed concentration with 100% positive reactions and %CV ≤35.

The linear interval of the assay was determined with methods based on the relative concentration approaches outlined in CLSI EP06 (4). During the analysis, the expected concentrations of each dilution were determined relative to the reported concentration of the most concentrated dilution. The relationship between reported and expected concentrations were modelled with a weighted least squares (WLS) regression. Predicted concentrations were calculated using this WLS model and the deviation between reported and predicted concentrations determined. The linear internal was considered the concentration range with absolute deviations ≤ 30%.

## Supplementary results

### Prototype Analytical Performance

The *P. aeruginosa* bacterial quantification assay (BQA) analytical performance were assessed including intra-assay precision, repeatability, LOD, LOQ and linearity (Table S1).

**Table S1:**
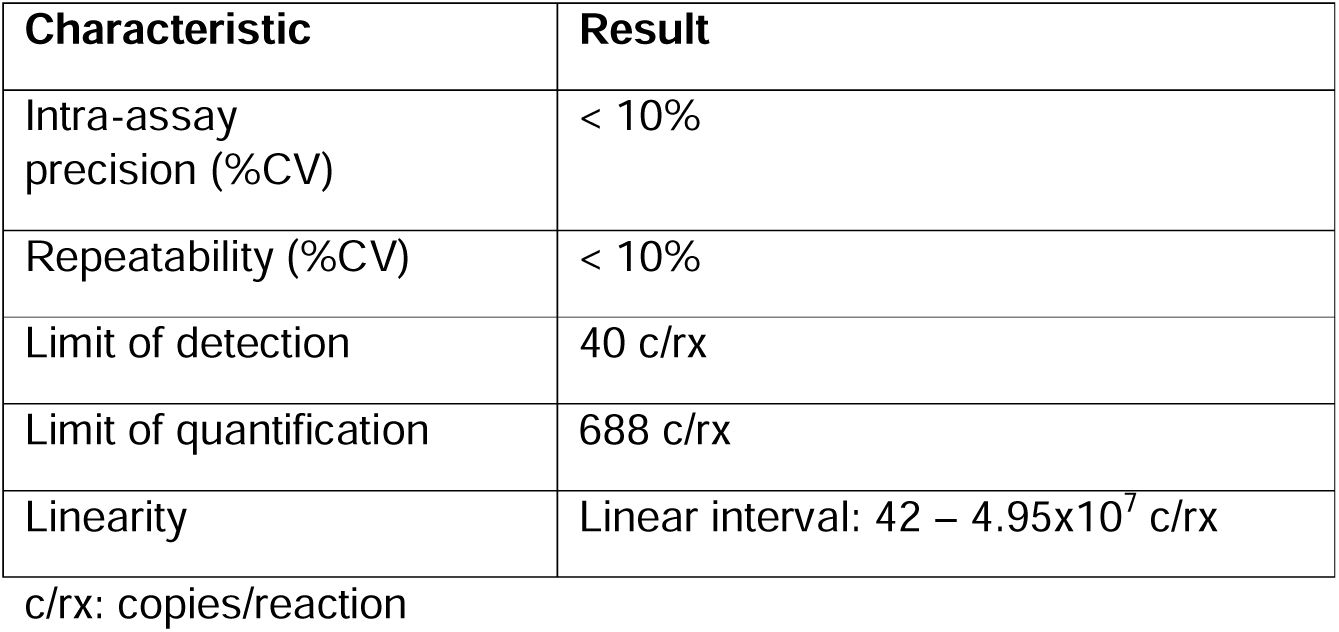
Summary of prototype analytical performance assessment.

| Characteristic | Result |
| --- | --- |
| Intra-assay precision (%CV) | < 10% |
| Repeatability (%CV) | < 10% |
| Limit of detection | 40 c/rx |
| Limit of quantification | 688 c/rx |
| Linearity | Linear interval: 42 – 4.95x10<sup>7</sup> c/rx |

c/rx: copies/reaction

**Table S2:** Detailed early detection analysis, including patient, status and quantification by rRNA and DNA.

| Patient | Status | PA CFU | rRNA Cq (mean) | DNA Cq (mean) | rRNA quantification (mean copies/reactions) | DNA quantification (mean copies/reactions) |
| --- | --- | --- | --- | --- | --- | --- |
| 1 | 298 days before culture positive | Neg | 27.96 | Neg | 8.66E+02 | Neg |
| 2 | 8 days before culture positive | Neg | Neg | Neg | Neg | Neg |
| 2 | 9 days before culture positive | Neg | Neg | Neg | Neg | Neg |
| 2 | 76 days before culture positive | Neg | Neg | Neg | Neg | Neg |
| 3 | 124 days before culture positive | Neg | Neg | Neg | Neg | Neg |
| 4 | 358 days before culture positive | Neg | Neg | Neg | Neg | Neg |
| 5 | 266 days before culture positive | Neg | 28.33 | 35.13 | 6.75E+02 | 9.98E+00 |
| 5 | 190 days before culture positive | Neg | Neg | Neg | Neg | Neg |
| 6 | 216 days before culture positive | Neg | Neg | Neg | Neg | Neg |
| 7 | 218 days before culture positive | Neg | Neg | Neg | Neg | Neg |
| 7 | 127 days before culture positive | Neg | 32.12 | Neg | 6.24E+01 | Neg |
| 7 | 268 days before culture positive | Neg | Neg | Neg | Neg | Neg |
| 8 | 63 days before culture positive | Neg | 32.06 | Neg | 5.47E+01 | neg |
| 9 | 266 days before culture positive | Neg | Neg | Neg | Neg | Neg |
| 10 | 137 days before culture positive | Neg | 17.94 | 26.26 | 7.28E+05 | 2.70E+03 |
| 11 | 129 days before culture positive | Neg | Neg | Neg | Neg | Neg |
| 12 | 41 days before culture positive | Neg | 27.65 | 33.51 | 1.07E+03 | 2.29E+01 |
| 13 | 484 days before culture positive | Neg | Neg | Neg | Neg | Neg |
| 14 | 67 days before culture positive | Neg | 23.56 | 28.11 | 1.66E+04 | 7.83E+02 |
| 15 | 93 days before culture positive | Neg | 22.95 | 25.86 | 2.52E+04 | 3.53E+03 |

**Table S3:**
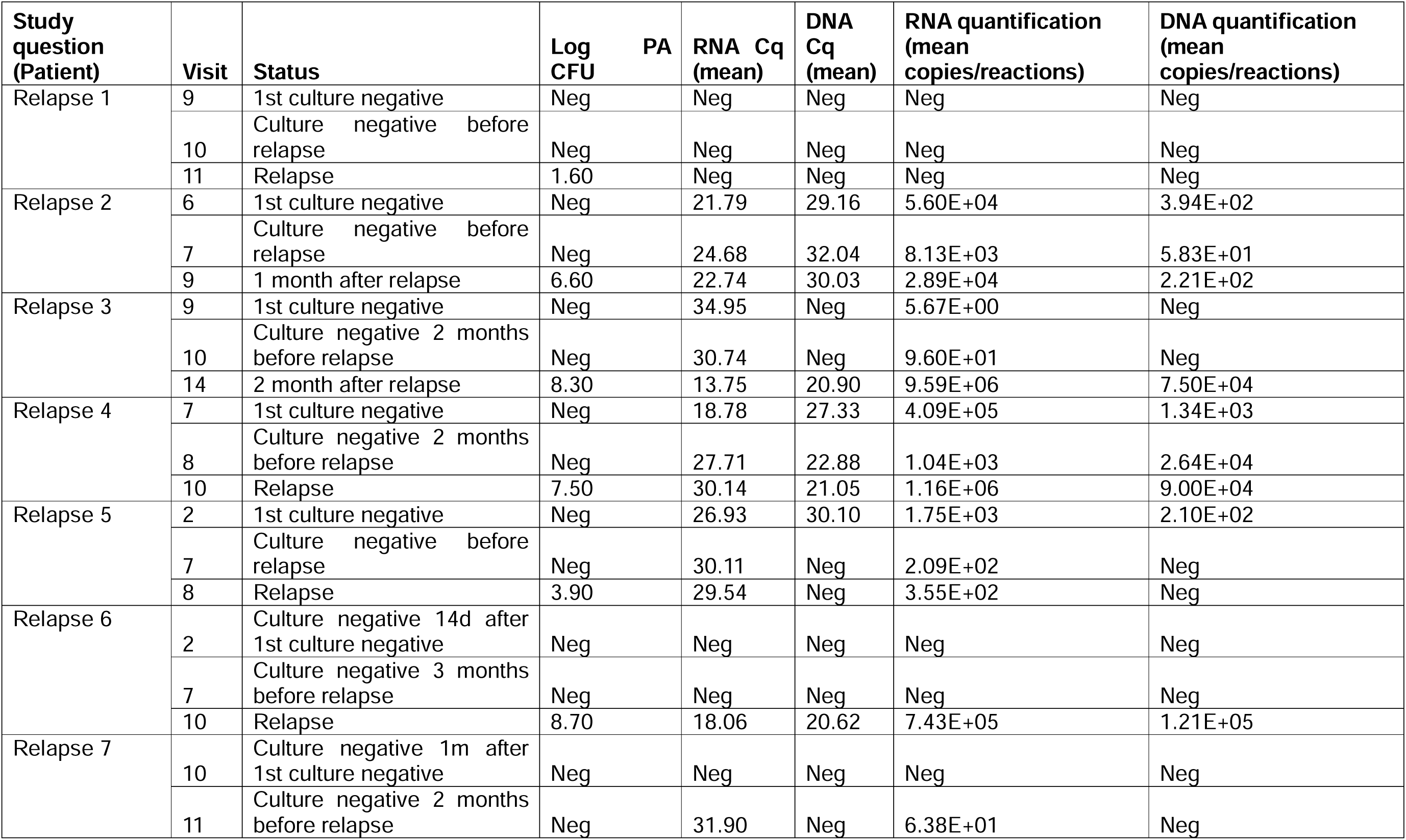
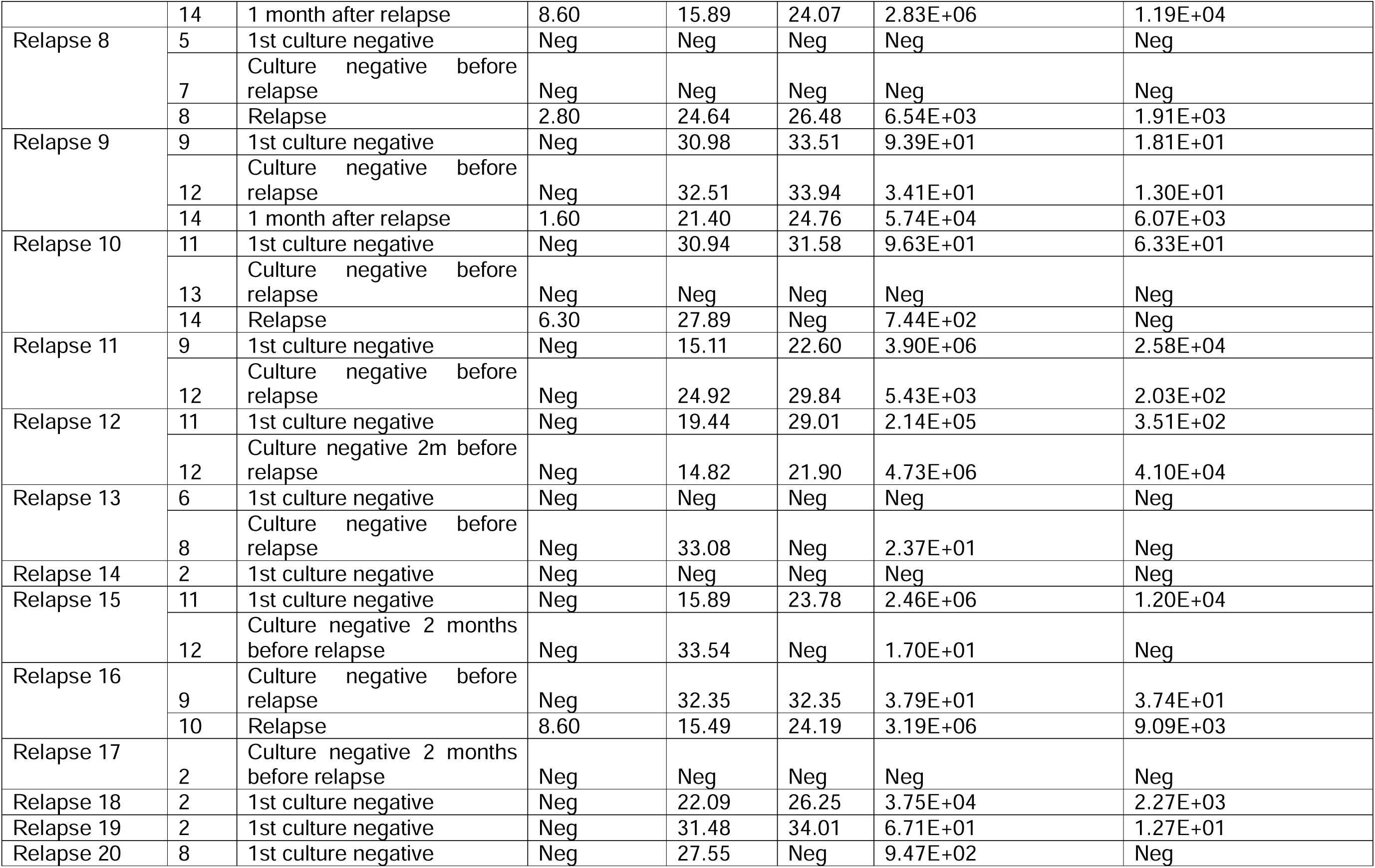
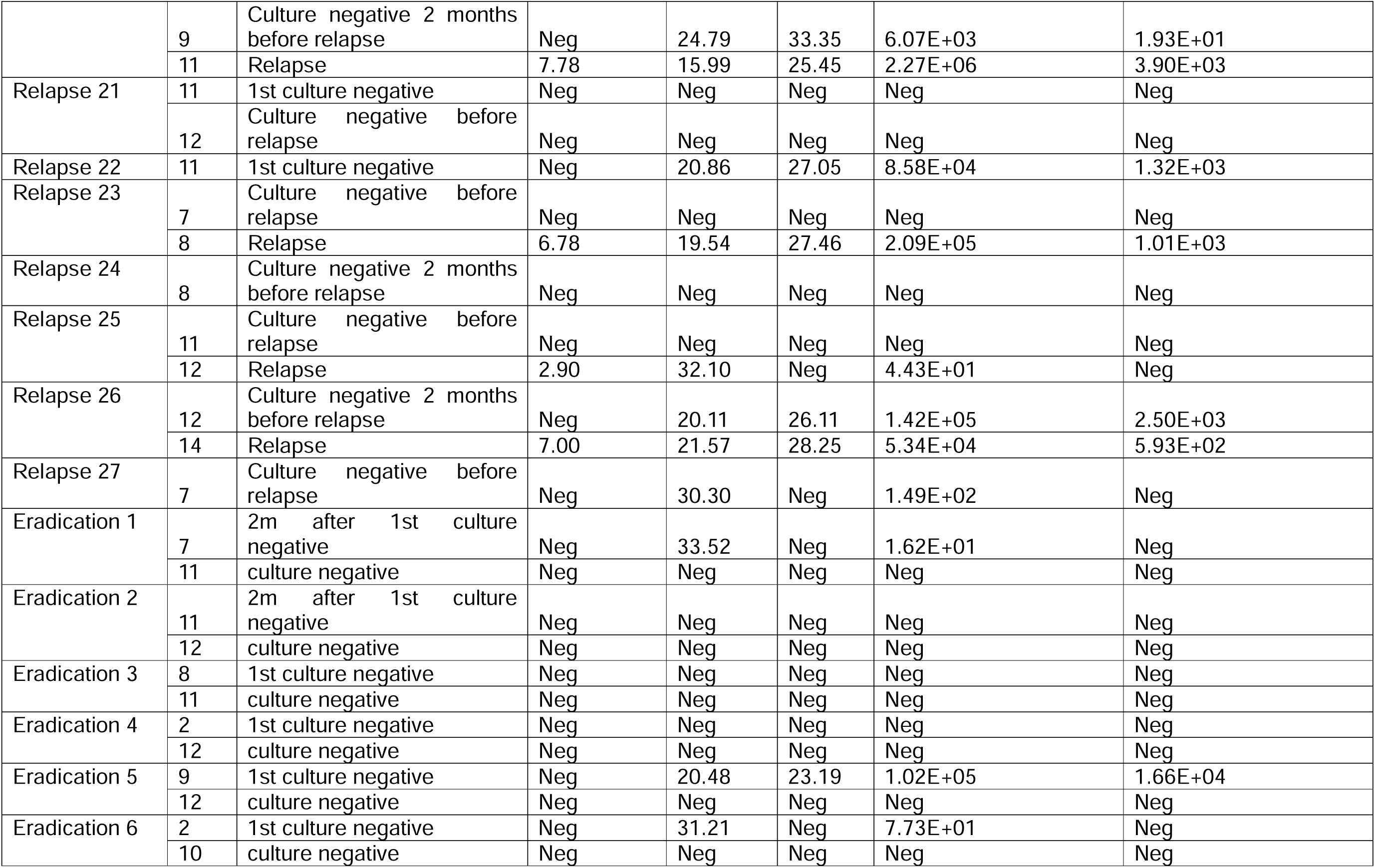
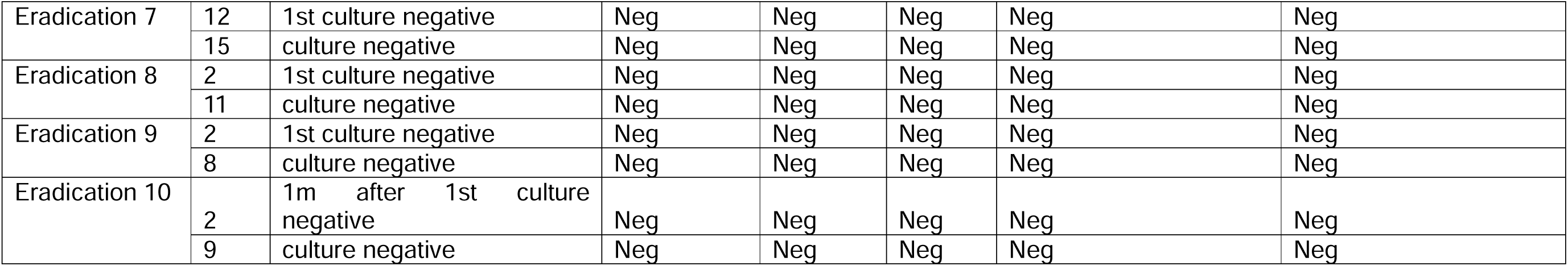
Detailed relapse and eradication analysis, including study question, patient, visit, status and quantification by rRNA and DNA.

## Notes

### Author Declarations

The study was approved by the research ethics committee in each participating country (London-Chelsea Research ethics committee, UK-18/LO/1935 (Coordinating country), Hannover Medical School, Germany, UZ Leuven, Belgium and UMC Amsterdam, Netherlands – ethical approval granted in all cases) and all patients gave written informed consent to participate.

